# Eye movement evaluation in Multiple Sclerosis and Parkinson’s Disease using a Standardized Oculomotor and Neuro-ophthalmic Disorder Assessment (SONDA)

**DOI:** 10.1101/2020.04.20.20072603

**Authors:** Alessandro Grillini, Remco J. Renken, Anne C. L. Vrijling, Joost Heutink, Frans W. Cornelissen

## Abstract

Evaluating the state of the oculomotor system of a patient is one of the fundamental tests done in neuro-ophthalmology. However, up to date, very few quantitative standardized tests of eye movements quality exist, limiting this assessment to confrontational tests reliant on subjective interpretation. Furthermore, quantitative tests relying on eye movement properties such as pursuit gain and saccade dynamics are often insufficient to capture the complexity of the underlying disorders and are often (too) long and tiring. In this study, we present SONDA (Standardised Oculomotor and Neurological Disorder Assessment): this test is based on analyzing eye tracking recorded during a short and intuitive continuous tracking task. We tested patients affected by Multiple Sclerosis (MS) and Parkinson’s Disease (PD) and find that: (1) the saccadic dynamics of the main sequence alone are not sufficient to separate patients from healthy controls; (2) the combination of spatio-temporal and statistical properties of saccades and saccadic dynamics enables an identification of oculomotor abnormalities in both MS and PD patients. We conclude that SONDA constitutes a powerful screening tool that allows an in-depth evaluation of (deviant) oculomotor behavior in a few minutes of non-invasive testing.

## 1 Introduction

Eye movements are a fundamental component of vision. Their evaluation is an important aspect of assessments in neurology and ophthalmology. Two neurological conditions in which oculomotor assessment has great clinical relevance are Multiple Sclerosis (MS) and Parkinson’s Disease (PD). In MS, the oculomotor assessment is relevant for diagnosis, for monitoring progress and for prognosis (Derwenskus et al., 2005; Elliot M. Frohman et al., 2005; Graves & Balcer, 2010), while in Parkinson’s Disease (PD), eye-movements can help in differentiating the “pure” form of PD from other forms of parkinsonism (Abadi & Gowen, 2004; Pretegiani & Optican, 2017; Rascol et al., 1991).

For both MS and PD, a confrontational assessment is the most commonly used technique for evaluating patients’ eye motility. While the simplicity of a confrontational assessment is attractive, its qualitative and subjective nature may leave subtle abnormalities undetected. In cases like sub-clinical manifestations of internuclear ophthalmoplegia in MS, or in PD, which typically exhibit eye movement disorders that are also present in the healthy elderly population, *quantitative* approaches are crucially needed (Marandi & Gazerani, 2019).

At present, quantitative assessments are relatively sparse and lack systematic, broadly accepted approaches. In pathological conditions, the oculomotor performance is usually parametrized using measures like the main-sequence (i.e. the relationship between saccadic amplitude and peak velocity (Bahill et al., 1975; Collewijn et al., 1988)), pursuit gain and onset latency, or generic positional errors, measured discretely by repeating many trials (Leigh & Zee, 2015). These trial-based approaches are often complex, repetitive and time-consuming, and - although being very suitable for research (for a noticeable example see DEMoNS (Nij Bijvank et al., 2018)) - they cannot be easily translated into daily clinical practice. Stressing the patients with such tests for longer than a few minutes can also exacerbate their symptoms: MS patients, for instance, are particularly affected by fatigue, which is reflected in changes in their saccadic peak velocity (Finke et al., 2012).

If we were to have a fast and simple yet powerful assessment of oculomotor quality, this could greatly benefit clinical practice. In this study we introduce SONDA, a Standardized Oculomotor and Neurological Disorders Assessment. This paradigm is partially based on the eye-movement cross-correlogram (Mulligan et al., 2013) applied to a continuous visual tracking task. This method measures several spatio-temporal properties (STP) of eye movements while patients track a moving dot with their eyes. In the analyses, the eye movements and their STP can be related to various aspects of visual functioning: e.g. the presence of visual field defects (Grillini et al., 2018) or spatial (Bonnen et al., 2015). The main advantage of this approach is that it provides a broad range of oculomotor measurements in a time-efficient and patient-friendly manner. Additionally, being able to provide an in-depth characterization of multiple aspects of eye movements is critical, because the clinical manifestations of oculomotor abnormalities are complex and variegated.

In MS, inflammatory demyelinating lesions in different brain areas result in a wide range of oculomotor disorders: most commonly, static and dynamic ocular misalignment (Serra et al., 2018) coupled with dysmetric saccadic behavior (Serra et al., 2003). Furthermore, fixation (Mallery et al., 2018), smooth pursuit (Lizak et al., 2016), and vestibulo-ocular responses (Huygen et al., 1986) are also often impaired. Most of these disorders are accentuated by the presence of internuclear ophthalmoplegia (INO). INO is a neuro-ophthalmic condition present in approximately one out of three MS patients (Jozefowicz-Korczynska et al., 2008) and is characterized by impaired adduction of conjugate lateral eye movements. The presence of INO is often determinant to convalidate the diagnosis of MS, especially if it is bilateral (Bolanos et al., 2004), but its sub-clinical manifestations are difficult to identify without quantitative approaches (T. C. Frohman et al., 2003).

Parkinson’s Disease (PD) and other forms of parkinsonism share some common features such as saccadic intrusions and square-wave jerks, but each clinical phenotype of parkinsonian disorders has a distinctive profile of oculomotor impairments (Abadi & Gowen, 2004; Pretegiani & Optican, 2017; Rascol et al., 1991). Eye-movement disorders typical of PD are impairment of self-paced saccades, saccadic hypometria, and saccadic fragmentation (Winograd-Gurvich et al., 2006): all contribute to disrupting smooth pursuit as well, due to the resulting inadequacy of catch-up saccades. These disorders, with the exception of the impairment of self-paced saccades, are also commonly present in otherwise healthy elderly individuals (Rottach et al., 1996). This implies that just finding an abnormality in eye movements will be insufficient to disentangle PD-related ones from those caused by normal aging, thus highlighting the need for multidimensional analysis of the oculomotor assessments.

We investigated whether the STP quantified with continuous gaze tracking is a sufficiently sensitive measure of the oculomotor disorders usually present in MS and PD. Furthermore, we extended the continuous gaze tracking method in order to measure saccadic dynamics continuously over time and to extract their frequency distribution. This will provide both a direct comparison between the eye-movement cross-correlogram STP and the saccadic main-sequence, as well as allowing the possibility of integrating these measurements into a single assessment.

More specifically, we answer the following research question: how well can the continuous gaze-tracking task of SONDA identify oculomotor abnormalities associated with MS and PD?

To summarize our findings, in both MS and PD patients, SONDA identified both marked and subtle abnormalities in oculomotor behavior based on only a few minutes of non-invasive testing.

Therefore, we conclude that SONDA allows an in-depth characterization of (deviant) oculomotor behavior and thus constitutes a powerful screening tool.

## 2 Methods

### 2.1 Observers

We assessed 71 participants divided into three groups. The first group comprised 50 healthy controls (age range 30-79, mean age 53±13.7, 26 females). The second group comprised 12 MS patients (age range 40-76, mean age 54±10.8, 8 females). The third group comprised 9 PD patients (age range 59-77, mean age 68±6.8, 2 females). All observers but one control and one MS patient had normal or corrected to normal visual acuity (below or equal to 0.10 LogMAR). All control participants were tested and screened with questionnaires to assess their familiarity with neurological and ophthalmological disorders at Royal Dutch Visio, Haren, The Netherlands. All observers gave their written informed consent prior to participation. The study was approved by the Medical Ethical Committee of the University Medical Center Groningen and the Ethics Committee of Psychology of the University of Groningen. The study followed the tenets of the Declaration of Helsinki.

### 2.2 Apparatus

A ‘Tobii T60 XL’ eye-tracker (Tobii Technology, Stockholm, Sweden) was used to record the eye movements of the observers. The recording was conducted binocularly, with a sampling frequency of 60 Hz. A custom-made 9-point calibration procedure was performed prior to each experimental session. The calibration was repeated until the average error was below 1 degree of visual angle and the maximum error below 2.5 deg. Stimuli were generated using custom-made scripts in MATLAB using the Psychophysics Toolbox (Brainard, 1997; Pelli, 1997) and displayed on the integrated screen of the Tobii T60 XL at a refresh rate of 60 Hz and viewed from a distance of 60 cm. Normally, this relatively low refresh rate is less suitable for the study of saccade dynamics, but we addressed this issue using a custom-made saccadic detection algorithm (see “Eye-tracking data analysis” paragraph). Head movements were minimized using a chin-rest with forehead support. All data were analyzed with custom made MATLAB scripts (R2017b, Mathworks). All display and analysis code is available upon request.

### 2.3 Continuous tracking task

The visual stimulus consists of a Gaussian blob of increased luminance moving along a random-walk trajectory on a uniform gray background (~ 140 cd/m2). The Gaussian blob is displayed at two contrast levels: at high contrast (50%) it has a peak luminance of ~385 cd/m2, while when presented at low contrast (10%), it has a peak luminance of ~160 cd/m2. Its full-width-at-half-maximum is 0.83 deg, roughly corresponding to the size III of a Goldman perimeter’s stimulus.

There were two stimulus conditions: in the *smooth pursuit* condition, the stimulus moved continuously along the random-walk path, while in the *saccadic pursuit* condition, an additional positional displacement to a random location on the screen was added to the trajectory, that occurred every two seconds. Each observer performed 6 trials of 20 seconds for each one of the conditions for a total of 12 trials and 4 minutes of total data acquisition. Short breaks in between trials were allowed. Additional detail regarding the visual stimulation can be found in Supplementary Material.

### 2.4 Eye-tracking data analysis

The extraction of the oculomotor spatio-temporal features is described in Supplementary Material.

Individual saccades were identified from the raw data using a customized MATLAB implementation based on the algorithm described by Behrens and colleagues. (Behrens et al., 2010) This algorithm allows saccade identification in eye movement time series using an adaptive threshold based on eye-movement acceleration. In short, the algorithm classifies any given time point at which the eye acceleration exceeds this adaptive threshold as being part of a saccadic movement. In their paper, the authors computed the threshold as a constant (K = 3.4) multiplied by the standard deviation of the acceleration distribution within a moving window of 200 frames, acquired at 1 kHz (i.e. a window of 0.2 sec). Our adapted version uses a moving window of 60 frames acquired at 60 Hz (i.e. a window of 1 sec). Empirically, we found our variant to be adequate for an accurate identification of saccades in our experiment. In contrast to Beherens and colleagues, we did not apply a low-pass filter to the velocity signals.

Since we conducted the gaze recording binocularly, we applied the saccade detection algorithm to the time series of each eye. An example of the saccades detected for one eye during one tracking trial is shown in Figure 1. Once the individual saccades are identified, their amplitude, peak velocity and direction are computed. Next, we used these parameters to compute the saccadic frequency and the saccadic dynamics distributions as a function of visual field eccentricity and direction. To determine whether the oculomotor behavior of patients differed from normal, we computed normative values based on the results of the control population. Because the age distribution of PD and MS patients differed, we computed separate normative values for each class of patients (see Results section, Figures 5-6, panels B-C)

**Figure 1.**
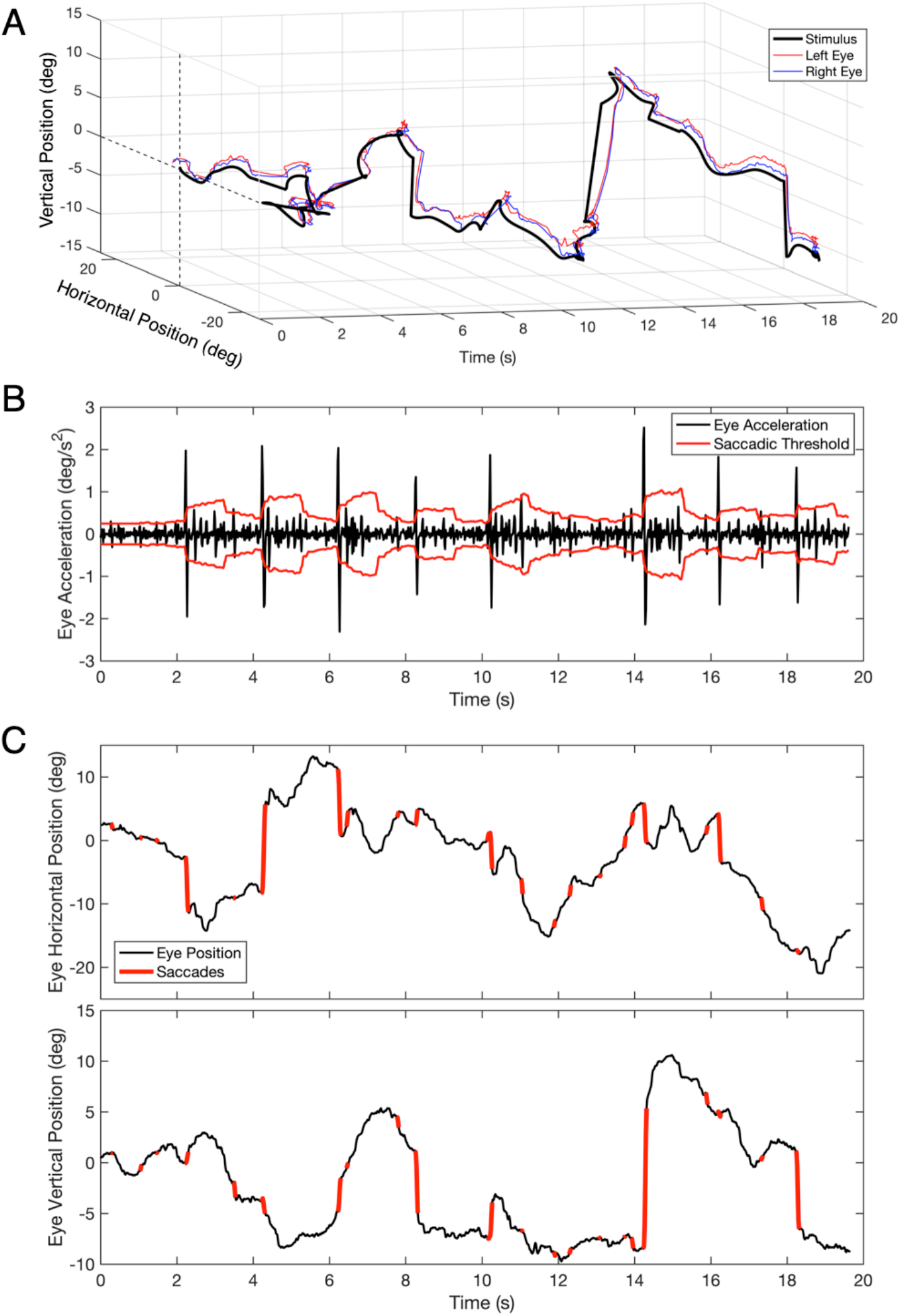
**A**. Example of a time series of eye movements positions recorded during a trial of 20 seconds of the tracking task. **B**. Example of eye acceleration and the value of the adaptive saccadic threshold for one eye plotted as a function of time. The adaptive threshold (in red) at any given time point is computed as the standard deviation of acceleration distribution in the preceding 60 samples, multiplied by a constant (K = 3.4). Whenever acceleration exceeds the momentary threshold, the time point is marked as being part of a saccade. **C**. Back-projection of the time points marked as being part of a saccade onto the original time series of eye positions (top figure showing the horizontal and the bottom figure showing the vertical component).

### 2.5 Statistical analysis

The results of the patients’ oculomotor behavior are expressed in modified z-scores computed as

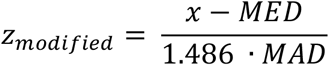

Where MED and MAD are the Median and Mean Absolute Deviation in the age-matched normative control population used as a reference. The individual saccades extracted with the algorithm described in the previous section, are binned in the function of their direction along the visual field, and used to display the saccadic frequency distribution (examples in panel B of Figures 5 & 6) and the saccadic dynamics (amplitude and peak velocity, examples in panel C of Figures 5 & 6).

For MS patients, the entire group of controls is used as normative data, as their age distributions are very similar (see Observers section). Since as a group the PD patients were significantly older, for the age-matched normalization it was necessary to create a subgroup of controls excluding all participants under 50 years of age. To take into account the different sizes of the groups (N_controls_ = 50, N_MS_ = 12, N_PD_ = 9), the statistical comparison between patients and controls have been performed using one-tailed Welch’s t-test for unequal variances with α = 0.05.

The analysis of oculomotor spatio-temporal properties (STP) resulted in 80 features (10 unique STP ✕ 2 axis ✕ 2 pursuit conditions ✕ 2 eyes). See Supplementary Material for a description of the individual spatio-temporal properties. For the purposes of the clustering analysis, we did not normalize the patients’ features with respect to the control group. Prior to performing the clustering analysis, we reduced the dimensionality of this dataset by employing a Principal Component Analysis (PCA) with the goal of removing redundant features. Based on the PCA, we identified the number of components sufficient to explain at least 95% of the variance. Following the PCA, we applied t-Distributed Stochastic Neighbor Embedding (tSNE) on the remaining components. This technique allows visualizing high-dimensional data by modeling each high-dimensional object as a two-(or three-) dimensional point in such a way that similar objects are modeled as nearby points and dissimilar objects are modeled as more distant points. Lastly, to identify clusters amongst the t-SNE points, we applied a k-means clustering algorithm. The appropriate number of clusters *k* was chosen using the Elbow Method (Thorndike, 1953) applied to the Within-clusters Sum of Squares (WSS). Based on the resulting *k*, the location and size of each of the clusters was computed.

## 3 Results

To summarize our findings, we find that the main-sequence alone is often insufficient to detect the presence of oculomotor abnormalities in either MS or PD. In contrast, we find that the STP obtained with our SONDA paradigm well separates PD patients from healthy controls. The STP of MS patients is insufficient to separate them from controls. Nevertheless, when we combine the STP with saccadic dynamics and frequency distributions, both PD and MS patients can be distinguished from controls.

### 3.1 Main-sequences of saccades in PD and MS do not differ from controls

First, we determined whether the oculomotor abnormalities of PD and MS patients are reflected in changes in the measures obtained with our continuous gaze tracking task. Figure 2 shows the main-sequence plots of amplitude vs. peak velocity for each saccade made by every patient and control made during the tracking task (algorithm described in Methods, Figure 1). We found no differences at the group level: both the amplitude and peak velocity distributions of the two patient groups overlap with those of the control group. For all three groups tested, we find an exponential relationship between amplitude and peak velocity. This behaviour is expected (Bahill et al., 1975). The prediction intervals (95%) of the patients’ groups data overlap with those computed for the control groups’ data.

**Figure 2.**
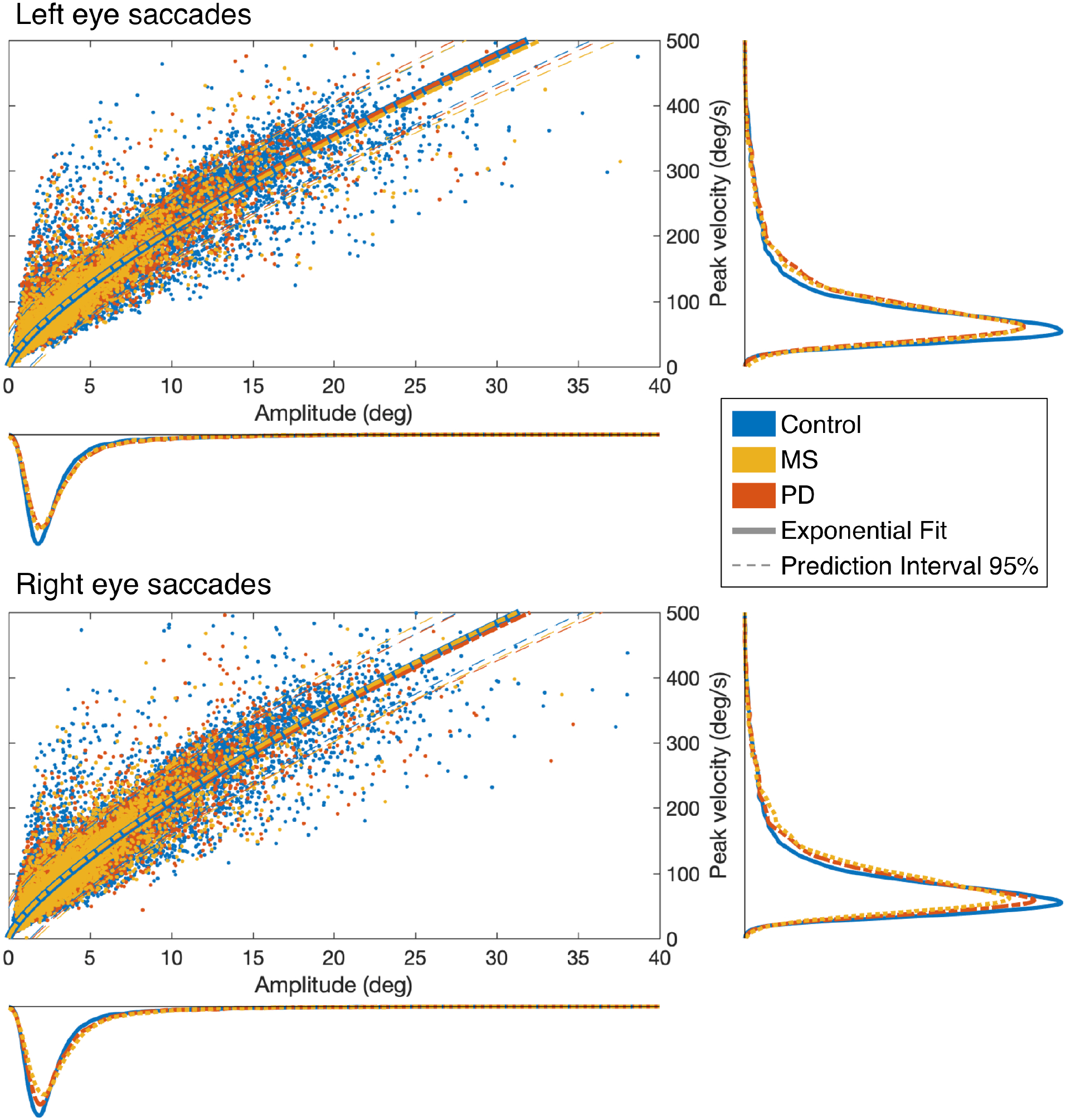
Saccadic main-sequences in the three examined groups of observers. The individual saccades (scatter plot) display the classic exponential relationship between amplitude and peak velocity, with no statistically significant differences between Control, MS and PD groups: all 95% prediction intervals overlap with each other. Also when taken separately (marginal histograms), the amplitude and peak velocity distributions do not differ between groups or between eyes.

### 3.2 Eye movement Spatio-Temporal Properties of PD and MS compared to normative data

Figure 3 shows the results of the analysis of the STP of eye movements made in the continuous tracking task.

**Figure 3.**
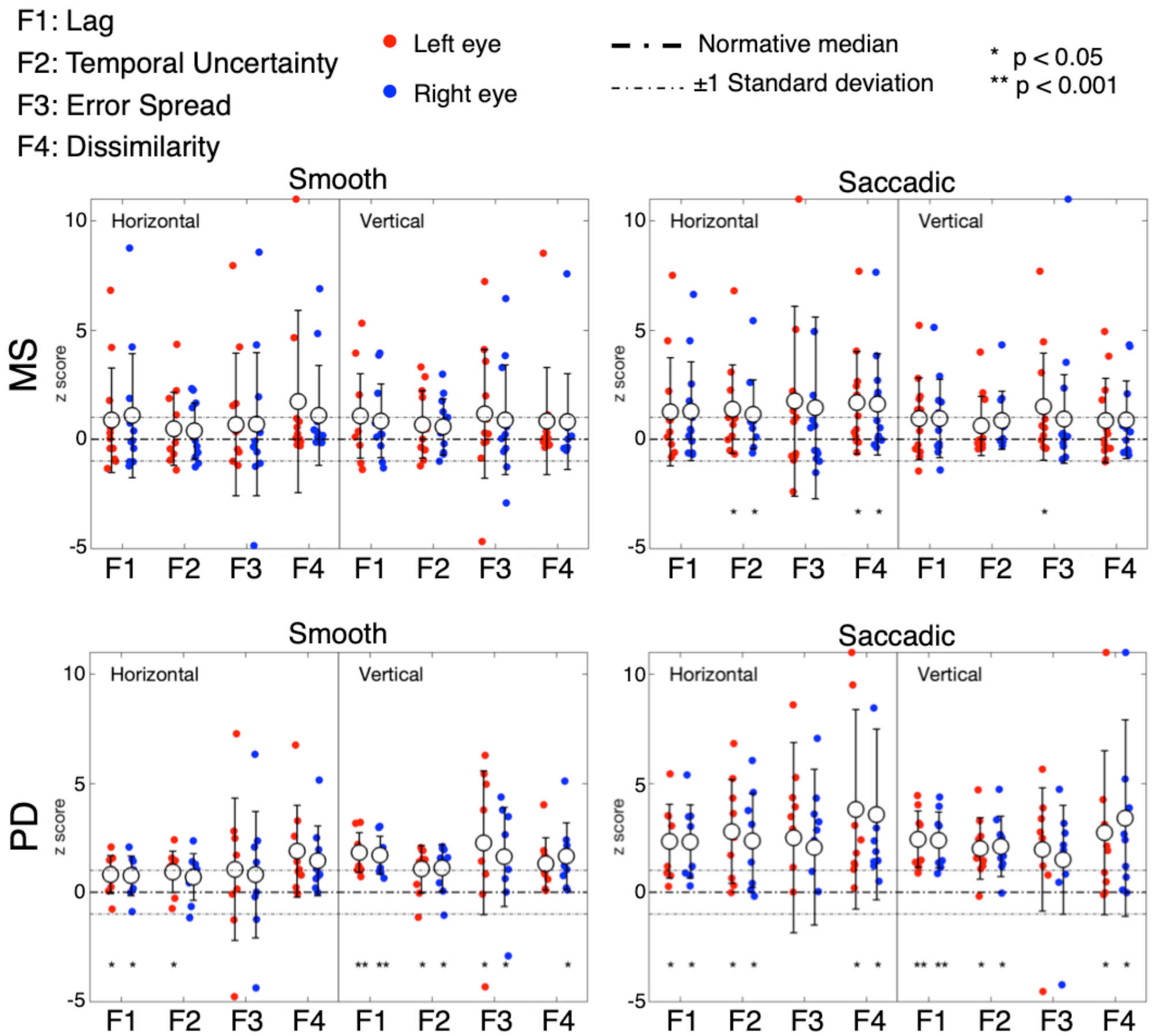
Oculomotor spatio-temporal properties in MS and PD cohorts, compared to an age-matched normative baseline. For the spatio-temporal analysis of the continuous gaze tracking task, we applied the eye-movement crosscorrelogram method (Grillini et al., 2018; Mulligan et al., 2013). From all features computed, we selected the two temporal and two spatial features which were shown to be most strongly modulated by stimulus velocity (Grillini et al., in preparation). The selected features are: “*lag”* (F1), *“temporal uncertainty”* (F2), *“error spread”* (F3) and “*dissimilarity”* (F4). “Lag” represents the delay between eyes and stimulus, measured as the temporal offset of the peak of the cross-correlation between stimulus and eye velocities. *“*Temporal uncertainty” represents the consistency of the velocity of the eye in relation to the velocity of the stimulus. “Error spread” is the standard deviation of the distribution of eye-stimulus positional deviations. “Dissimilarity” is the inverse of the cosine similarity computed between the arrays of gaze coordinates and stimulus coordinates.

In MS patients (upper row), we find a substantial overlap of their features with the normative ones. We can observe a higher variability compared to controls and PD (lower row), especially in the Smooth condition. Statistically significant differences (indicated by an asterisk) are found for the temporal uncertainty and the movement dissimilarity features, but only in the Saccadic condition, where participants had to make saccades in order to track the rapid, random, displacements of the target.

In PD patients (lower row), nearly all features are significantly different from the normative ones. The magnitude and variance of the differences are larger in the Saccadic condition, similar to what we found in the MS patients.

For both MS and PD patients, there were neither significant differences for the left and right eyes, nor for the horizontal and vertical components of the eye movements.

These results indicate that MS patients had abnormalities primarily in saccades, while PD patients had abnormalities in both saccades and smooth-pursuit eye movements.

### 3.3 Classification of the neurological disorder based on oculomotor abnormalities

We examined whether it is possible to identify the underlying neurological conditions of individual patients based exclusively on the eye-movements measurement. The results are shown in Figure 4. Of the 80 initial features (10 oculomotor properties ✕ 2 axis ✕ 2 pursuit conditions ✕ 2 eyes), the PCA indicated that 11 components are sufficient to explain 95% of the variance in the data (Figure 4-A). We then used the t-SNE algorithm to embed these 11 principal components into two final components for clustering and visual representation. We found that the optimal number of clusters to represent the data is five (Figure 4-B). The results of the k-mean analysis are shown in Figure 4-C. The leftmost cluster [1] contains nearly all PD patients and two of the MS patients. Being most separated from the other clusters, this cluster appears to group the cases with the most severe oculomotor impairments. The rightmost cluster [2] includes the vast majority of control subjects, thus grouping those without an oculomotor impairment. It partially overlaps with clusters [3] and [4] which contain participants from various groups and together cover nearly all MS patients. These clusters, therefore, appear to group patients with a suspected impairment of different degrees of severity. The remaining cluster [5] comprises outliers which, upon verification, were associated with very noisy data and/or exceptionally poor performance. Altogether, these results show that despite the relatively low number of subjects, a classification based exclusively on STP is reasonably effective for Parkinson’s Disease but ineffective for MS.

**Figure 4.**
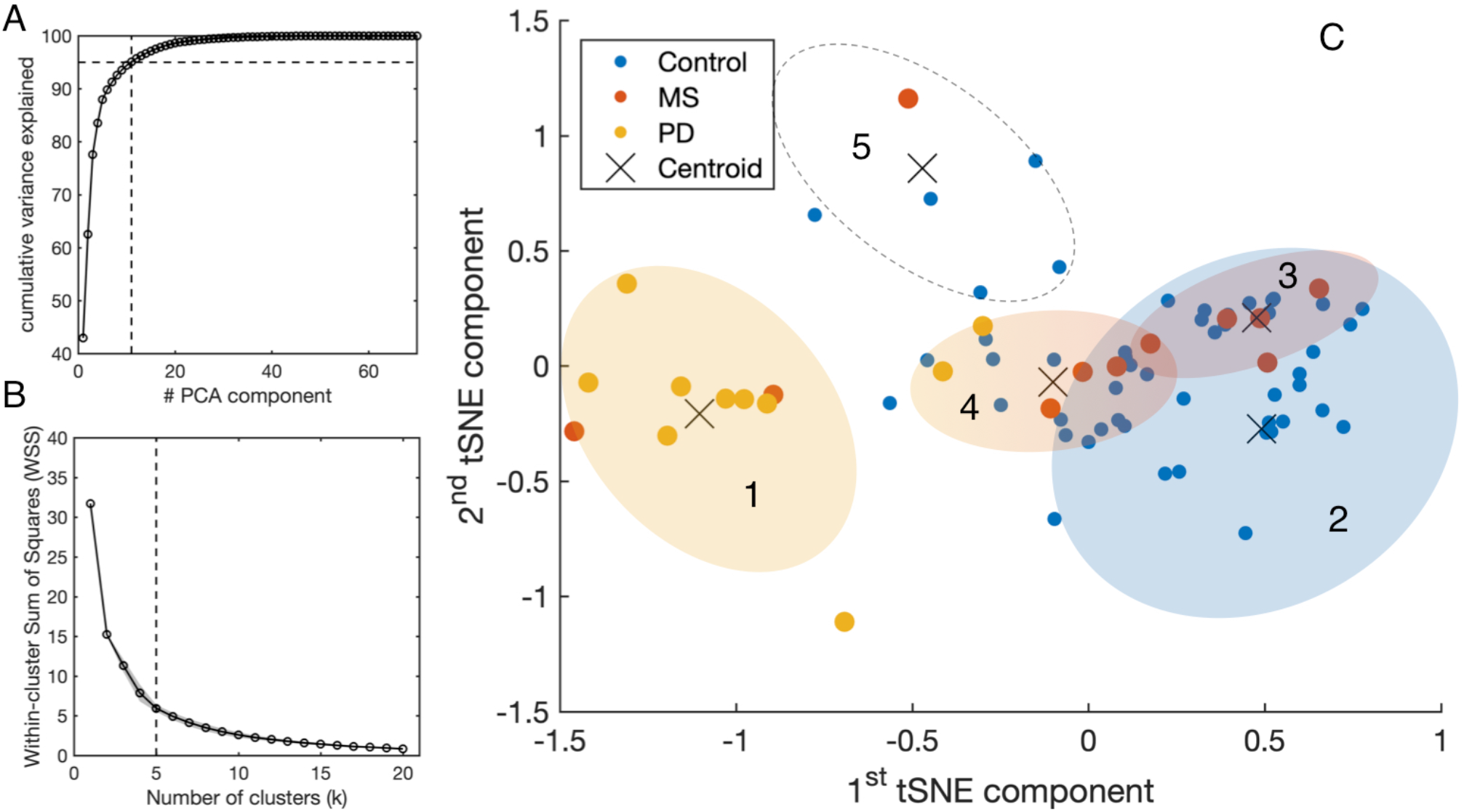
Results of STP-based clustering analysis for the entire tested population. All the spatio-temporal features available from SONDA were used as an input for the PCA (see Supplementary Material), and the resulting components were processed with tSNE to represent the high-dimensionality dataset into a lower-dimensionality space for clustering purposes. The number of clusters has been computed using the “elbow method” with the Within-clusters Sum of Squares (WSS) as a parameter. We then used the k-means clustering algorithm (*k* = 5) applied to the outcome components of t-SNE.

**Figure 5 & 6.**
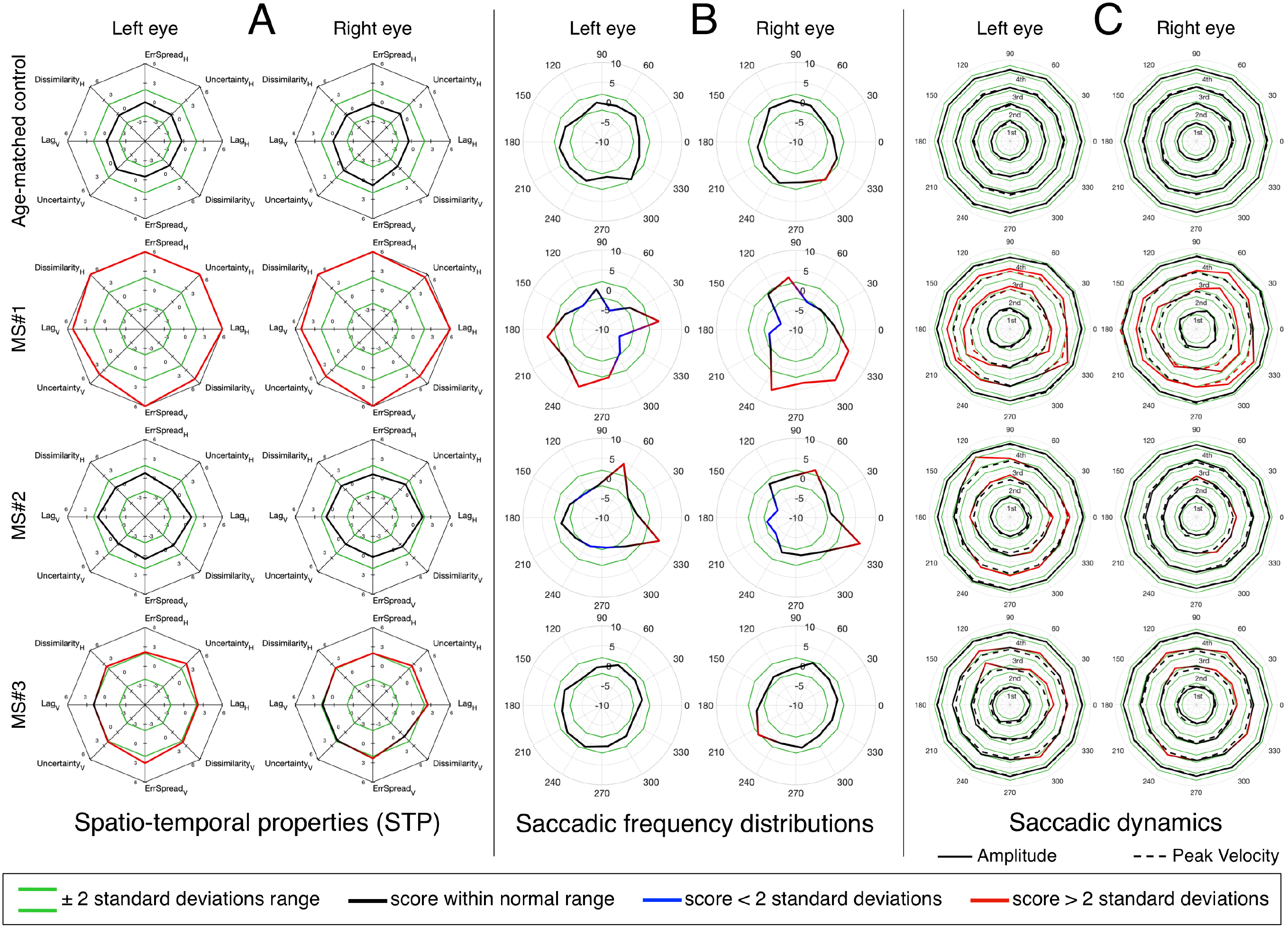
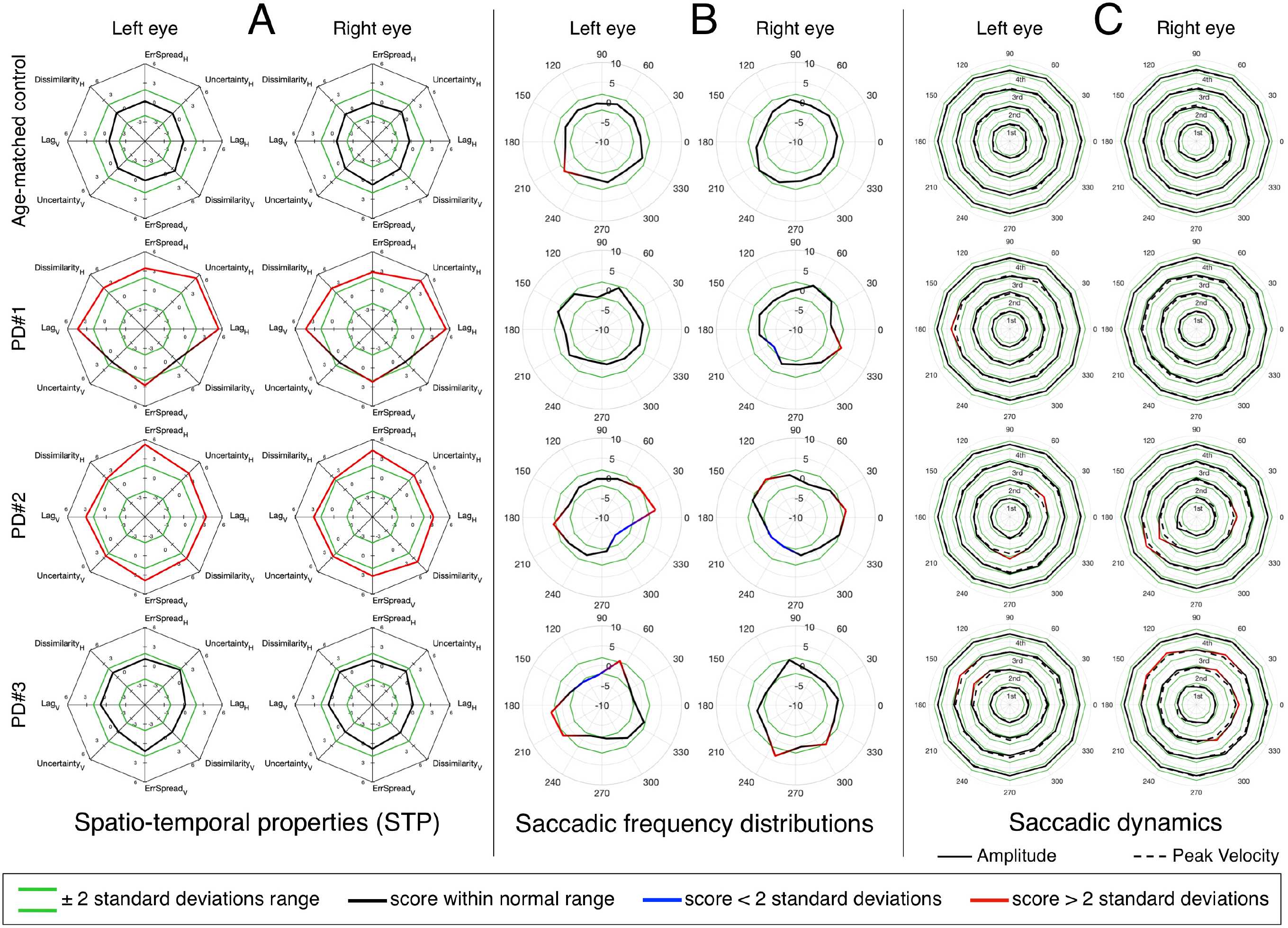
An in-depth overview of the oculomotor performance of three MS patients, three PD patients, and two age-matched controls. **A**. Normalized spatio-temporal properties (STP). We consider an STP feature abnormal if its deviates 2 or more standard deviations from the normative value (indicated in green). **B**. Normalized saccadic frequency distributions. **C**. Normalized saccadic dynamics (amplitude and peak velocity), as a function of saccade direction. The results are stratified per quartile, separately for amplitude and peak velocity. Each quartile has its own normative boundaries.

### 3.4 Combination of STP with statistical and dynamic properties of saccades

Next, we verified whether taking into account the frequency distribution of saccades and their dynamic properties as a function of direction (i.e. visual field polar angle), would allow for a better discrimination between the underlying disorders. Figure 5 and 6 show the results for three MS and three PD patients respectively, and two age-matched control observers. From each patient group, we selected three patients that showed either mixed results regarding the STP, the saccadic frequency and saccadic dynamics, or that had an additional diagnosis (e.g. INO in addition to MS). The results for the left and right eye are displayed separately.

Panel A shows part of the STP previously presented in Figure 3. We present the features for the horizontal and vertical components of the *saccadic pursuit* condition, which we already showed to be the condition with the most significant differences. Our results show that the STP and the saccadic dynamics are not always in agreement and thus convey different types of information that may be relevant for a clinical interpretation. Patient MS#1 had been previously diagnosed with internuclear ophthalmoplegia (INO) and has severely impaired STP (Fig. 5-A), a dysmetric saccadic behavior with reduced adduction frequency (Fig. 5-B), and an abnormal range of saccadic amplitudes and peak velocities of which both are hypermetric (Fig. 5-C). Patient MS#2 has normal STP (Fig. 5-A), an anisotropic saccadic frequency distribution skewed rightward and upward binocularly (Fig. 5-B). The saccades made by both eyes are in the normal velocity range, but their amplitude is mildly hypermetric, especially along a near vertical and in a horizontal rightward direction (Fig. 5-C). Patient MS#3 has moderately impaired STP (Fig. 5-A), a normal saccade frequency distribution (Fig. 5-B), and hypermetric amplitude of the vertical saccades, with a normal peak velocity for both eyes (Fig. 5-C).

Patient PD#1 has impaired STP, with a strong oculomotor lag in both horizontal and vertical components (Fig. 6-A), while both their saccadic frequency distribution (Fig. 6-B) and saccadic dynamics (Fig. 6-C) are within the normative range. Patient PD#2 shows severely impaired STP (Fig. 6-A), an unusual “diagonally skewed” saccadic frequency distribution for both eyes (with the upward component slanted towards the nasal side; Fig. 6-B) and the saccadic peak velocity within the normative range and nearly perfect normal saccadic amplitude (Fig. 6-C). Finally, patient PD#3 has normal STP (Fig. 6-As) and shows saccadic frequency distributions that deviate from normal (Fig. 6-B) and show slightly hypermetric vertical amplitude of the diagonal saccades. The peak velocities for both eyes were normal (Fig. 6-C).

## 4 Discussion

The main contribution of the present study is the introduction of a novel method to perform a time-efficient (less than 5 minutes), non-invasive, neuro-ophthalmic screening that is based on an in-depth characterization of oculomotor behavior in response to a simple visual tracking task.

First, our results indicate that the most commonly used conventional measures such as saccadic amplitude and peak velocity by themselves, are often insufficient to fully characterize the oculomotor behavior of PD and MS patients. Moreover, we show that the computation of the spatio-temporal properties (STP) of eye-movements recorded during a continuous tracking task can help detect abnormalities in the oculomotor behavior. Therefore, we conclude that a combination of improved conventional (saccadic frequency distribution and dynamics) and our entirely new spatio-temporal analyses are an effective screening tool to comprehensively assess the oculomotor behavior of a patient. The clinical relevance of this will be discussed below for each class of patients.

### 4.1 SONDA identifies oculomotor abnormalities in Multiple Sclerosis

The parameters of the main-sequences - saccadic amplitude and saccadic peak velocity - of MS patients showed an almost identical distribution to those of the controls (see Figure 2). This is consistent with the literature which indicates that most CNS lesions of this disease have not been associated with unique identifiable clinical symptoms in eye-movements (Elliot M. Frohman et al., 2005). Thus far, the majority of the studies on oculomotor abnormalities in MS patients focused on the detection of internuclear ophthalmoplegia (INO). This disorder may be present (often subclinically) in ~30% of MS patients and is usually identified by measuring the versional dysconjugacy index (VDI), which is the ratio between abducting and adducting eye saccadic dynamics (E. M. Frohman et al., 2002; Jozefowicz-Korczynska et al., 2008; Nij Bijvank et al., 2019). However, despite that relying on the main-sequence parameters alone is insufficient, standardized screening tests capable of detecting other manifestations of oculomotor abnormalities in MS do not exist.

With our novel “SONDA” approach introduced in this study, in MS we found preserved smooth pursuit responses and two abnormal features in the saccadic pursuit condition (see Figure 3). This is consistent with the notion that the most common saccadic issue in MS is saccadic dysmetria (Serra et al., 2003), present especially in patients with demyelination affecting the cerebellar peduncles (E. M. Frohman et al., 2001). However, given the relatively small sample (NMS = 12) the significant difference seems to be driven mostly by a few severely impaired patients, rather than a general decrease in performance.

An interesting result emerged from the clustering analysis shown in Figure 4. In this case, we performed a multivariate analysis based on the whole feature-space of STPs obtained with SONDA, rather than selecting a few parameters a priori. We found that MS patients, although not well separable from controls, nevertheless tended to cluster (clusters [3] and [4]) based solely on their STPs. This suggests the presence of subtle oculomotor abnormalities that require more detailed quantification of the eye movements.

To further explore this hypothesis we also quantified per eye the saccadic frequency and the saccadic dynamics distributions (Figures 5 and 6). These extended analyses revealed the presence of abnormalities even in patients falling within the normative STP ranges. For instance, patient MS#2 has a normal STP (panel 5A), only slightly abnormal saccadic dynamics (panel 5C), but a very peculiar saccadic frequency distribution (panel 5B). This latter analysis revealed an abnormal prevalence of horizontal rightward saccades for both eyes. This is analogous to what has been observed in pathological nystagmus: a slow-drift of eccentric gaze in one direction followed by a resetting saccade in the other, which is caused by a failure of the common neural integrator of eye movements in the brainstem (Elliot M. Frohman et al., 2005). While the other two MS patients (MS#1 and MS#3) have an abnormal STP (panel 5A). A further inspection of their saccadic frequency distributions does differentiate between them: MS#3 has a normal saccadic frequency distribution (panel 5B) and only slightly hypermetric vertical saccades (panel 5C). The oculomotor abnormalities of patient MS#3 might have gone unnoticed if the assessment had been limited to the saccadic dynamics. On the other hand, besides an abnormal STP, patient MS#1 also displays overall hypermetric saccades, as well as the suppression of adducting saccades (panel 5B) that is typical of INO.

### 4.2 SONDA identifies oculomotor abnormalities in Parkinson’s Disease

In PD, slower saccades should be expected in advanced cases (Rottach et al., 1996; Vidailhet et al., 1994; White et al., 1983). Yet, at the group level, the prediction intervals for the saccadic main-sequence of the PD patients overlap with those of the control group (see Figure 2). In contrast a significant difference is found in all examined STP, except for the spread of the positional error (Figure 3). Noticeably, we found marked differences along the vertical axis of the smooth pursuit condition for almost the entire group, although mild cases of PD are expected to be similar to age-matched control observers (Rottach et al., 1996; Waterston et al., 1996). This can be due to the fact that while unpredictable pursuit should be poor in PD, the predictive component of pursuit is generally preserved (Gilaie-Dotan et al., 2013). However, our stimulus trajectory is completely unpredictable (by design it does not contain any autocorrelation; see Supplementary Material). This explains why our approach was sensitive to PD related abnormalities.

Regarding saccades, one of the characteristic oculomotor disorders in PD is the impairment of self-generated saccades with the relative sparing of visually-driven saccades, which only deteriorate as the disease progresses (Leigh & Zee, 2015). In our tracking task we tested only visually-driven saccades but still found significant differences in the STP of most patients. This is important, because both at the group-level (Figure 2) and at the single-subject level (Figure 6C) the saccadic dynamics of the PD patients were remarkably normal. It suggests that SONDA is a sensitive paradigm.

For the PD patients, the clustering analysis based on SONDA’s STP resulted in the separation of most PD individuals from the controls (Figure 4). This is quite remarkable considering that, for pure Parkinson’s Disease (which all patients in our group were) the eye movements usually only show minor disturbances that are common in healthy elderly subjects as well (Leigh & Zee, 2015). Note that this contrasts with other forms of parkinsonism. Furthermore, one of the main oculomotor abnormalities in PD is the difficulty in generating internally driven saccades. Most errors are made when PD patients are required to switch between instructions (e.g. switching between pro- and antisaccades (Cameron et al., 2012; Rivaud-Pechoux et al., 2006; Terao et al., 2013)). Such switching is not demanded by our paradigm. Nonetheless, abnormal oculomotor behavior could be clearly identified in all PD patients.

An in-depth inspection of the oculomotor behavior of single patients (Figure 6) showed that, analogously to MS, an analysis of the saccadic frequency distribution can reveal subtle abnormalities that other measures (STP and saccadic dynamics) cannot (see Figure 6, PD#3 for an example). Nevertheless, in the majority of the PD patients, the STP by itself was sufficient to identify eye movement abnormality (as already revealed by the clustering analysis).

### 4.3 SONDA is clinically relevant

The SONDA paradigm allows for a comprehensive examination of eye-movements that provides detailed information on the dynamics and statistical properties of saccades, as well as a novel set of spatio-temporal properties that quantify both smooth and saccadic pursuit behavior. Given the large number of parameters, our technique also opens up possibilities for machine-learning-aided diagnosis of neuro-ophthalmic and neurological impairment based on eye movements. We and others have previously demonstrated the feasibility of such an approach for identifying (simulated) ophthalmic disorders (Crabb et al., 2014; Grillini et al., 2018), and for neurological conditions (the latter based on participants viewing natural scenes (Tseng et al., 2013)). Compared to other high-throughput eye-tracking tests, SONDA has two main advantages. First, excluding calibration (that is common for all eye-tracking tests), the whole assessment takes only 4 minutes (compared to ~20 minutes for the approach of Tseng et al (Tseng et al., 2013) and ~25 minutes for DEMoNS (Nij Bijvank et al., 2018)). Second, our task and stimulus were designed to be as intuitive and simple as possible to avoid confounding factors that may be caused by natural or more complex stimuli, or complicated instructions resulting in a higher cognitive load.

### 4.4 Future applications

Objective, quantitative eye-movement-based tests are increasingly becoming an important aid in diagnosing and monitoring neuro-ophthalmic conditions of which the oculomotor manifestations cannot always be clinically detected with confrontational assessments (Bedell & Stevenson, 2013). For instance, in some patients oscillopsia occurs in conjunction with eye movements that are too small to be detected using an ophthalmoscope, yet that are large enough to cause visual complaints (Smith, 1986). Another example of the use of eye-movement-based metrics is the assessment of surgical or pharmacological interventions for nystagmus (Hertle et al., 2003; McLean et al., 2007). Such disorders are rather common in MS when lesions involve the brainstem and the cerebellum (Elliot M. Frohman et al., 2005) and can occur in PD as well, sometimes causing oscillopsia (Kaski & Bronstein, 2017).

Besides MS and PD, the new parameters provided by SONDA can be relevant in the evaluation of other neurological and psychiatric disorders such as mild traumatic brain injury (Thiagarajan et al., 2011), schizophrenia (Holzman et al., 1973) or dementia with Lewy bodies (Mosimann et al., 2005). In these disorders, the parameters of the saccadic main-sequence of patients often overlap with those of controls. For this reason, outcomes cannot be considered definitively diagnostic (Serra et al., 2003). A multi-dimensional characterization of oculomotor properties, such as provided by SONDA, could thus also be beneficial in these conditions.

### 4.5 Limitations

Our present study still has some limitations that should be addressed in future follow-up. First, the number of observers tested in comparison to the number of features analyzed was only just enough to use t-SNE and k-mean clustering (N = 71 observers and 11 PCA components; see Figure 4). Combined with the imbalance within groups (N_control_ = 50, N_MS_ = 12, N_PD_ = 9) this may have prevented an optimal separation of the MS patients from the controls. Another aspect that requires further investigation is the sensitivity/specificity of the measures that were expressed as a function of the visual field polar angle (Figures 5 & 6, panels B & C). The number of bins used to determine the polar histograms was chosen arbitrarily and might affect the false negative/positive rate depending on the data available in each bin.

### 4.6 Conclusions

We presented SONDA, a novel paradigm to assess oculomotor abnormalities that can be linked to neurological disorders. We tested patients affected by Multiple Sclerosis and Parkinson’s Disease and showed that: (1) the parameters of the saccadic main-sequence alone are insufficient to separate patients from healthy controls; (2) spatio-temporal and statistical properties of the eye movements obtained with SONDA, when combined with detailed analysis of saccadic dynamics allow for a correct identification of the oculomotor abnormalities and identification of the underlying neurological disorder. Our test constitutes an attractive approach for quantitative neuro-ophthalmic screening, being non-invasive, intuitive and fast to administer.

## Data Availability

Data is available upon request to the corresponding author.

## 5 Author Contributions

AG, AV and FC designed the experiment; AG and RR analyzed the data; AG and FC wrote the manuscript; AV, JH and FC wrote the grant; all authors reviewed the manuscript.

## 6 Conflict of interest

The authors AG and RR are listed as inventors on the European patent application “**Grillini, A**., Hernández-García, A., **Renken, J. R**. (2019). Method, system and computer program product for mapping a visual field. EP19209204.7” which is partially based on the content of the Supplementary Material of this manuscript.

The author AG is currently a majority shareholder of Reperio B.V. (Dutch chamber of commerce registration: 76175596), but was not at the time the research was conducted.

The authors AV, JH and FC declare that the research was conducted in the absence of any commercial or financial relationships that could be construed as a potential conflict of interest.

## 7 Funding

This project has received funding from the Programmaraad Visuele Sector (grant agreement “STIP-MS”), the European Union’s Horizon 2020 research and innovation programme under the Marie Sklodowska-Curie (grant agreement No 641805 “NextGenVis”) and the Graduate School Medical Sciences (GSMS) of the University Medical Center Groningen.

## 8 Acknowledgments

We are thankful to Nadine Naumann and Teatske Hoekstra from Royal Dutch Visio for their support in assessing patients.

